# Prevalence and Risk Factors for Non-Communicable Chronic Diseases in Low and Middle-Income Countries: A Systematic Review and Meta-Analysis

**DOI:** 10.1101/2024.10.15.24315536

**Authors:** Sancho Pedro Xavier, Ana Raquel Manuel Gotine, Melsequisete Daniel Vasco, Audêncio Victor

**Affiliations:** Institute of Collective Health, Federal University of Mato Grosso. Av. Fernando Correa da Costa, n° 2367 - Bairro Boa Esperança. Cuiabá - MT - 78060-900, Brazil; USP, School of Public Health, University of São Paulo (USP), Avenida Doutor Arnaldo, 715, São Paulo, São Paulo, 01246904, Brazil; Institute of Collective Health, Federal University of Bahia. R. Basílio da Gama s/n, Canela. 40110-040 Salvador, BA, Brazil

**Keywords:** non-communicable chronic diseases, Hypertension, Diabetes, Systematic review, Meta-analysis, Meta-regression, Low- and middle-income countries (LMIC)

## Abstract

**Background:** Non-communicable chronic diseases (NCDs) have become increasingly prominent in low- and middle-income countries (LMIC), driven by a rapid rise in their incidence. Current estimates suggest that these conditions account for approximately 80% of deaths in these regions. This study aimed to analyze the prevalence of NCDs and their associated risk factors in LMIC.

**Methods:** Electronic searches were conducted in the PubMed, Embase, Scopus, Cochrane, and Virtual Health Library (VHL) databases between June and July 2023. Studies on the prevalence of NCDs, with or without associated risk factor analysis, were included. The quality of these studies was assessed using NIH tools, and a meta-analysis was conducted using the random-effects model.

**Results:** A total of 34 studies on hypertension and 22 studies on diabetes were included in the systematic review and meta-analysis. The estimated prevalence of hypertension was 24% (95% CI: 21.0; 28.0) and diabetes mellitus (DM) was 11% (95% CI: 10.0; 13.0), with future predictions for similar populations ranging from 11.0-46.0% for hypertension and 6.0-21.0% for DM. Geographic analysis revealed a lower prevalence of hypertension in Latin America and the Caribbean (7.0%) with no statistically significant differences compared to other regions (p-value = 0.101). The prevalence of DM was lower in Sub-Saharan Africa (5.0%; p-value < 0.001). The identified risk factors for hypertension included increased age, male sex, elevated BMI, alcohol consumption, excessive salt intake, and stress. For diabetes, the risk factors were increased age, lack of religious affiliation, elevated BMI, family history of DM, hypertension, high hemoglobin concentration (HbA1c), waist-to-hip ratio, smoking, and infection with Taenia spp.

**Conclusion:** NCDs such as hypertension and DM pose a growing public health challenge in low- and middle-income countries. Our findings may assist policymakers in identifying high-risk groups and recommending appropriate prevention strategies.

**Systematic Review Registration:** The protocol was submitted for registration with the International Prospective Register of Systematic Reviews (PROSPERO) (registration number: CRD42024520601).

## Introduction

Non-communicable diseases (NCDs) have received increasing attention in low- and middle-income countries (LMIC) [1]. They are responsible for approximately 35 million deaths, representing 60% of all global deaths, with 80% occurring in these countries [2]. Hypertension and diabetes mellitus (DM), two of the main NCDs, are among the most fatal and prevalent conditions in the adult population, constituting a significant public health threat [3], which are impacted by urban and industrial growth and are often associated with unhealthy lifestyles, such as tobacco and alcohol consumption, physical inactivity, and inadequate diets [4, 5]. NCDs have ceased to be an emerging problem in developing countries, assuming an alarming dimension and reaching epidemic proportions [2]. It is estimated that by 2030, eight of the ten leading causes of death will be related to these conditions [6]. The impact of NCDs has serious implications for global social and economic development, particularly for LMIC [6]. Critical risk factors include tobacco consumption, unhealthy diets, physical inactivity, and excessive alcohol consumption [2].

Although there is substantial research on the prevalence and associated risk factors of NCDs in low- and middle-income countries, the results of individual studies are often insufficient to guide clinical decisions, practical actions, or public health policies. Therefore, systematic reviews are essential as they critically evaluate all available evidence and combine the results to offer more robust conclusions on the subject [7]. Thus, this research aimed to conduct a comprehensive systematic review and meta-analysis of the existing literature to provide a clearer understanding of the magnitude of these diseases and their risk factors, offering greater consistency and validity to previous findings. We hope that the findings of this research will contribute to the development of effective public health strategies, including prevention, detection, treatment, and control, to reduce morbidity and mortality associated with these conditions and their complications.

## Methods

### Study protocol

The identification of records, title, and abstract screening, as well as the evaluation of the eligibility of full texts for inclusion in the final analysis, were carried out according to the PRISMA guidelines (Preferred Reporting Items for Systematic Reviews and Meta-Analyses) [8]. This study was registered in PROSPERO under the reference number CRD42024520601, available at: https://www.crd.york.ac.uk/prospero/display_record.php?ID=CRD42024520601.

### Search strategy

Potentially eligible studies were identified through a systematic search of the PubMed, Embase, Scopus, Cochrane, and Virtual Health Library (VHL) databases. Initially, studies published up to June 2023 were included. An updated search was conducted in September 2024 to include studies published between June 2023 and September 2024. This update ensured that the most recent and relevant studies were incorporated into our review. The search terms used during the research were: (“(Prevalence trends” OR “prevalence patterns” OR “prevalence rates” OR “prevalence changes”) AND (“non-communicable chronic diseases” OR “non-communicable disorders” OR “non-communicable conditions” OR “NCDs”) AND (“low- and middle-income countries” OR “LMICs” OR “developing countries” OR “emerging economies”) AND (“risk factors” OR “determinants” OR “contributors” OR “influencing factors”), based on the POT principles (P - Population of low- and middle-income countries; O - prevalence and risk factors for non-communicable chronic diseases; T - Observational studies) to retrieve relevant articles through the databases mentioned above. The research aimed to find observational and interventional studies reporting the prevalence of non-communicable chronic diseases and their risk factors. No language or publication date restrictions were applied.

### Study selection

Three study team members (ARG, MV, and SX) independently reviewed all titles and abstracts after the initial removal of duplicates, using Rayyan [9], and applied the following inclusion criteria: (i) observational studies that collected primary data through questionnaires, interviews, physical exams, or other data collection methods; (ii) inclusion of data on prevalence and/or risk factors; (iii) studies focused on adult populations (18 years or older) diagnosed with NCDs; and (iv) studies that address populations from countries classified as LMIC according to the World Bank classification. Studies with a small sample size (due to high selection bias), studies focused on pediatric populations, institutions, or specific groups such as women exclusively, patients under care including pregnant women, conference papers or abstracts, articles without full text, and studies whose data could not be obtained from corresponding authors were excluded. Two team members (SX and AV) read and evaluated the full texts of the remaining articles, resolving disagreements through discussions until a consensus was reached.

### Study Outcomes

The primary outcomes of interest were the prevalence of hypertension and DM. Hypertension was defined as elevated and persistent blood pressure, with systolic blood pressure (SBP) ≥ 130 mmHg and/or diastolic blood pressure (DBP) ≥ 80 mmHg or reported use of antihypertensive medication [3]. Fasting plasma glucose (FPG) (≥7.0) mmol/l (126 mg/dl) was classified as DM [3, 10, 11]. Additionally, participants who were taking prescribed medications to lower their elevated blood glucose levels were classified as having DM [3].

### Data extraction and risk of bias assessment

Data from eligible studies were extracted by one reviewer into an Excel spreadsheet template suggested in a study [7], with a second reviewer checking each cell. To ensure the compliance of extracted data, a review was conducted by all members, and finally, a lead reviewer went through each cell. The extracted data included the last name of the first author, year of publication, journal, Region (country), study design, Inclusion criteria, Population of the study, Sample size, follow-up duration, and disease-associated factors.

The NIH Study Quality Assessment Tools for observational and cross-sectional studies were used to assess the quality of the included studies (see S1 in supplements). Generally, the tool includes 14 questions to assess the quality of the articles, with categorical responses (Yes (1), No (0), Other (CD, NR, NA), where CD means cannot determine; NA, not applicable; NR, not reported). The overall score was calculated by summing the scores of all items, with “yes” equating to one, while “no” and “NA” equated to zero. Each article was assigned a score to classify them as poor, fair, or good studies, with a score of 0-5 considered poor, 6-9 considered fair, and 10-14 considered good [7].

### Data analysis

A random-effects model (DerSimonian & Laird) was used for the meta-analysis due to the assumed heterogeneity among the studies. The I² statistic [12] and the Q test [13] were used to assess heterogeneity among the studies. The I² index refers to the proportion of observed variance, and 25%, 50%, and 75% or more of the statistics indicated low, medium, and high heterogeneity, respectively [14]. Publication bias was assessed using a visual inspection of the symmetry of the funnel plot, followed by the application of Egger’s test [15] and Begg’s rank test [16] for confirmation. P-values less than 0.05 indicated evidence of publication bias among the included studies. Subgroup analysis was conducted according to study design (cohort and cross-sectional), publication year (2012-2014, 2015-2019, and 2020-2022), sample size, geographic region, and study quality (Fair, Good, and Poor). However, as these assessments do not provide information about the sources of heterogeneity, meta-regression analysis was applied to explore potential sources of heterogeneity in the combined estimates. Additionally, a leave-one-out sensitivity analysis was performed by iteratively removing one study at a time to examine the effects of a single study on the overall estimate [17]. The results were presented in a forest plot as a point estimate with 95% confidence intervals (95% CI). Pooled Odds Ratios (ORs) were used to describe the possible association between outcomes and predictors. A p-value < 0.05 was considered statistically significant. All analyses were performed using R (R Foundation for Statistical Computing) version 4.4.1, utilizing the meta package.

## Results

### Characteristics of included studies

A total of 1264 potential studies were identified: 969 articles from PubMed, 267 from Cochrane, 9 from Scopus, 5 from VHL, and 4 from Embase. Figure 1 shows the search results and the reasons for exclusion during the article selection process. A total of 38 articles published between 2011 and 2024 were included to analyze the prevalence of Hypertension (in 34 studies) and DM (in 22 studies) in LMIC, of which 16 articles [3–5, 10, 11, 18–31] were used to analyze the factors associated with the prevalence of these diseases. The characteristics of the included studies are summarized in **Table 1**. According to the study design, of the included studies, 32 were cross-sectional studies and 6 were cohort studies. Among these, 22 articles were from Sub-Saharan Africa, 12 from South Asia, 5 from the Middle East and North Africa, 4 from East Asia and the Pacific, and 1 from Latin America and the Caribbean. In this meta-analysis, 923,251 individuals were involved in the analysis of hypertension prevalence, and 322,513 for diabetes, with 163,577 and 41,100 having hypertension and DM, respectively. Regarding study quality, the scores ranged from 6 to 13 **(see S2 in supplements).**

**Fig. 1.**
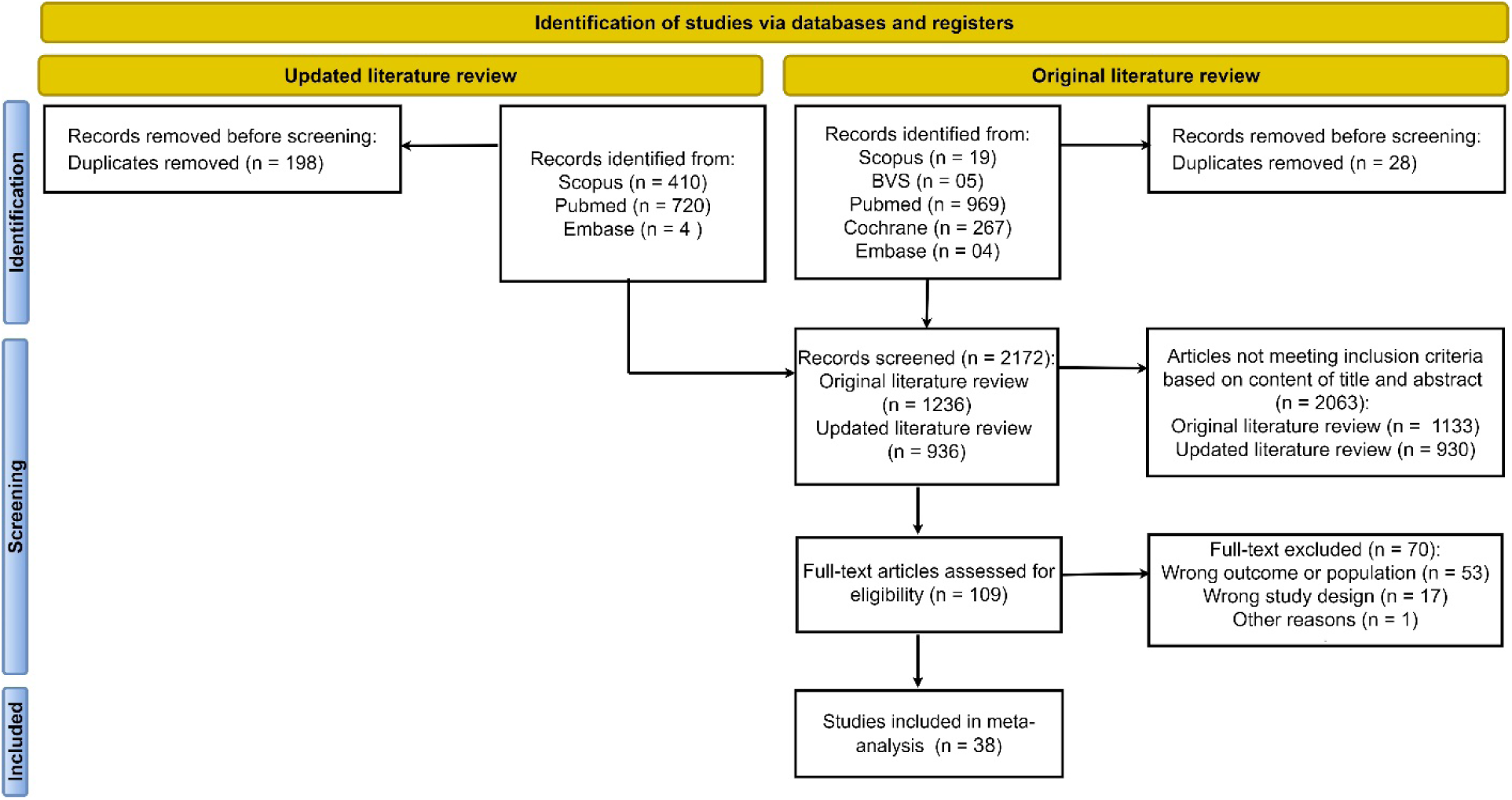
Flowchart for finding studies

**Fig.2.**
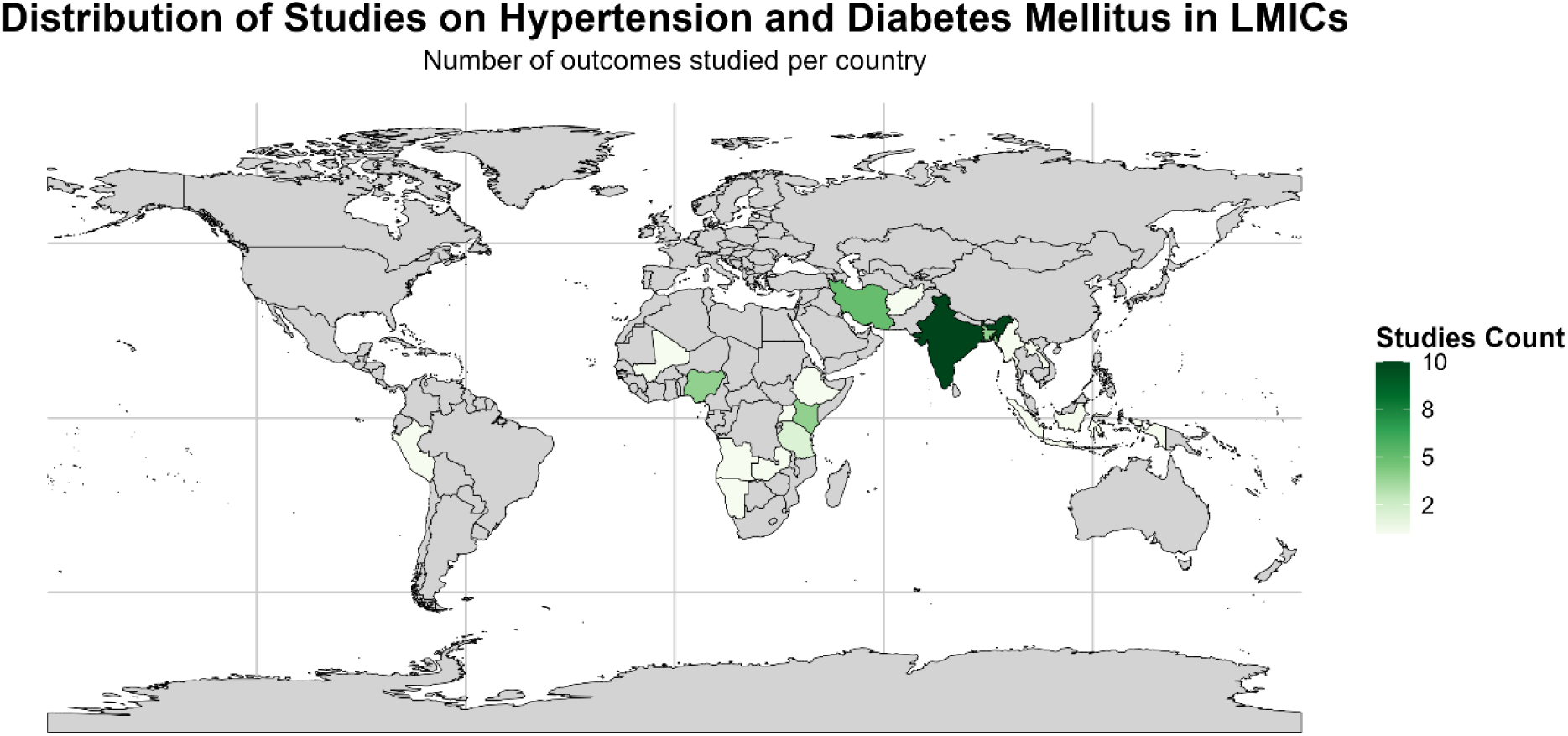
Distribution of the countries included in the studies

**Table 1.** Baseline characteristics of all the studies included in the meta-analysis.

|  |  |  |  |  |  |  |  |  |  |
| --- | --- | --- | --- | --- | --- | --- | --- | --- | --- |
| **Authors** | **Year** | **Country** | **Study Design** | **Sample size** | **Mean/Median Age** | **Age included** | **Outcome** | **Prevalence** | **SEP\*** |
| Vellakkal et al. 2013 [71] | 2013 | India | Cross-sectional | 12198 | Not reported | 18 or + | Hypertension | 19.0 | 0,004 |
| Rahman, 2022 [3] | 2022 | Bangladesh | Cross-sectional | 12290 | 31 | 18 or + | Hypertension and Diabetes | 36.98; 11.29 | 0.004; 0.003 |
| Diawara et al. 2023 [27] | 2023 | Mali | Cross-sectional | 412 | Not reported | 20 or + | Diabetes | 7.5 | 0.013 |
| Giri et al. 2022 [28] | 2022 | India | Cross-sectional | 819 | Not reported | 18 or + | Hypertension | 16.7 | 0.013 |
| Safarpour et al. 2022 [72] | 2022 | Iran | Cohort | 4997 | 48.41 | 35 or + | Hypertension and Diabetes | 18.63; 15.67 | 0.006; 0.005 |
| Kumma et al. 2021 [10] | 2021 | Etiopia | Cross-sectional | 2486 | 35 | 25 or + | Hypertension | 31.3 | 0.009 |
| Khamseh et al. 2021 [11] | 2021 | Iran | Cohort | 163770 | Not reported | 35 or + | Diabetes | 15.0 | 0.001 |
| Simmons et al. 2021 [73] | 2021 | Bangladesh | Cross-sectional | 8019 | Not reported | 18 or + | Hypertension | 21.2 | 0.005 |
| Kiani et al. 2021 [74] | 2021 | Iran | Cohort | 7978 | 49.48 | 35 or + | Hypertension and Diabetes | 27.1; 12.0 | 0.005; 0.004 |
| Khoo et al. 2022 [75] | 2021 | Malasia | Cross-sectional | 34149 | Not reported | 18 or + | Hypertension | 18.0 | 0.002 |
| Sivanantha et al. 2021 [5] | 2021 | India | Cross-sectional | 2415 | 44.3 | 18 or + | Hypertension and Diabetes | 33.6; 26.7 | 0.000; 0.009 |
| Riaz BK et al. 2020 [4] | 2020 | Bangladesh | Cross-sectional | 8185 | Not reported | 18 or + | Hypertension | 21.0 | 0.005 |
| Okello et al. 2020 [76] | 2020 | Kenya, Nigeria, Tanzania, and Uganda | Cross-sectional | 3549 | 39.7 | 18 or + | Hypertension | 25.4 | 0.007 |
| Ongosi et al. 2020 [29] | 2020 | Kenya | Cross-sectional | 593 | Not reported | 25 or + | Hypertension and Diabetes | 26.2; 7.7 | 0.02; 0.01 |
| Alsaud et al. 2020 [77] | 2020 | Jordanians | Cross-sectional | 1449 | 44.35 | 20 or + | Hypertension and Diabetes | 37.5; 21.5 | 0.012; 0.011 |
| Odili et al. 2020 [78] | 2020 | Nigeria | Cross-sectional | 4192 | 46.7 | 18 or + | Hypertension | 38.1 | 0.007 |
| Mirzaei et al. 2020 [79] | 2020 | Iran | Cohort | 8749 | Not reported | 20 or + | Hypertension and Diabetes | 18.6; 14.1 | 0.004; 0.004 |
| Thakur et al. 2019 [30] | 2019 | India | Cross-sectional | 5078 | Not reported | 18 or + | Hypertension and Diabetes | 26.2; 15.5 | 0.006; 0.005 |
| Latt et al. 2019 [80] | 2019 | Myanmar | Cross-sectional | 8757 | Not reported | 25 or + | Diabetes | 10.5 | 0.003 |
| Htun et al. 2018 [31] | 2018 | Laos | Cross-sectional | 1528 | 54.9 | 35 or + | Diabetes | 22.8 | 0.011 |
| Peltzer and Pengpid 2018 [81] | 2018 | Indonesia | Cross-sectional | 29965 | 43.3 | 18 or + | Hypertension | 33.4 | 0.003 |
| Demisse et al. 2017 [18] | 2017 | India | Cross-sectional | 3227 | 41.1 | 18 or + | Hypertension | 27.4 | 0.008 |
| Thakur et al. 2016 [32] | 2016 | India | Cross-sectional | 5127 | Not reported | 18 or + | Hypertension and Diabetes | 40.1; 14.0 | 0.007; 0.005 |
| Seclen et al. 2015 [82] | 2015 | Peru | Cross-sectional | 1677 | Not reported | 25 or + | Diabetes | 7.0 | 0.006 |
| Talaei et al. 2014 [1] | 2014 | Iran | Cohort | 3283 | 50.7 | 35 or + | Hypertension and Diabetes | 22.5; 10.4 | 0,007; 0,005 |
| Romdhane et al. 2014 [19] | 2014 | Tunisia | Cross-sectional | 7700 | 49.0 | 35 or + | Diabetes | 15.1 | 0.004 |
| Shah and Afzal 2013 [83] | 2013 | India | Cross-sectional | 1768 | Not reported | 20 or + | Hypertension and Diabetes | 18.16; 16.63 | 0.009; 0.009 |
| Ayah et al. 2013 [20] | 2013 | Kenya | Cross-sectional | 2061 | 33.4 | 18 or + | Diabetis | 3.2 | 0.004 |
| Daniel et al. 2013 [84] | 2013 | Nigeria | Cross-sectional | 964 | 38.41 | 20 or + | Hypertension | 38.2 | 0.02 |
| Pires et al. 2013 [21] | 2013 | Angola | Cross-sectional | 1464 | 33.7 | 18 or + | Hypertension | 23.0 | 0.01 |
| Muyer et al. 2012 [85] | 2012 | Congo | Cross-sectional | 1759 | 51.5 | 20 or + | Hypertension and Diabetes | 9.8; 4.8 | 0.009; 0.005 |
| Hendriks et al. 2012 [86] | 2012 | Nigeria, Kenya, Tanzania, Namibia | Cross-sectional | 2678, 2111, 1046, 1733 | 45.3, 40.9, 36.8, 36.9 | 18 or + | Hypertension | 21.0, 20.2, 19.0, 32.0 | 0.007, 0.009, 0.01, 0.01 |
| Nsakashalo-Senkwe et al. 2011 [22] | 2011 | Zambia | Cross-sectional | 1928 | Not reported | 25 or + | Diabetes | 4.0 | 0.004 |
| Anjana et al. 2023 [87] |  | India | Cross-sectional | 113043 | Nao reportado | 20 or + | Diabetes | 7.3 | 0.001 |
| Dadras et al. 2024 [23] | 2024 | Afghanistan | Cross-sectional | 3890 | 35.0 | 18 or + | Diabetes | 11.0 | 0.005 |
| Seenappa et al. 2024 [24] | 2024 | India | Cross-sectional | 743067 | Nao reportado | 18 or + | Hypertension | 15.9 | 0.000 |
| Bhatia et al. 2023 [25] | 2023 | India | Cohort | 3183 | Nao reportado | 45 or + | Hypertension | 20.8 | 0.007 |
| Kibria et al. 2024 [26] | 2024 | Bangladesh | Cross_sectional | 7932 | 39.2 | 18 or + | Diabetes | 9.9 | 0.003 |

## Meta-analysis

### Prevalence of hypertension and diabetes

According **to Figure 3**, 34 studies on the prevalence of hypertension were included in the global analysis of this meta-analysis. The estimated combined proportion of hypertension, obtained through the random-effects model, was 0.24 (95% CI: 0.21; 0.28). The prediction for future populations with similar characteristics ranged between 0.09 and 0.49.

**Fig.3.**
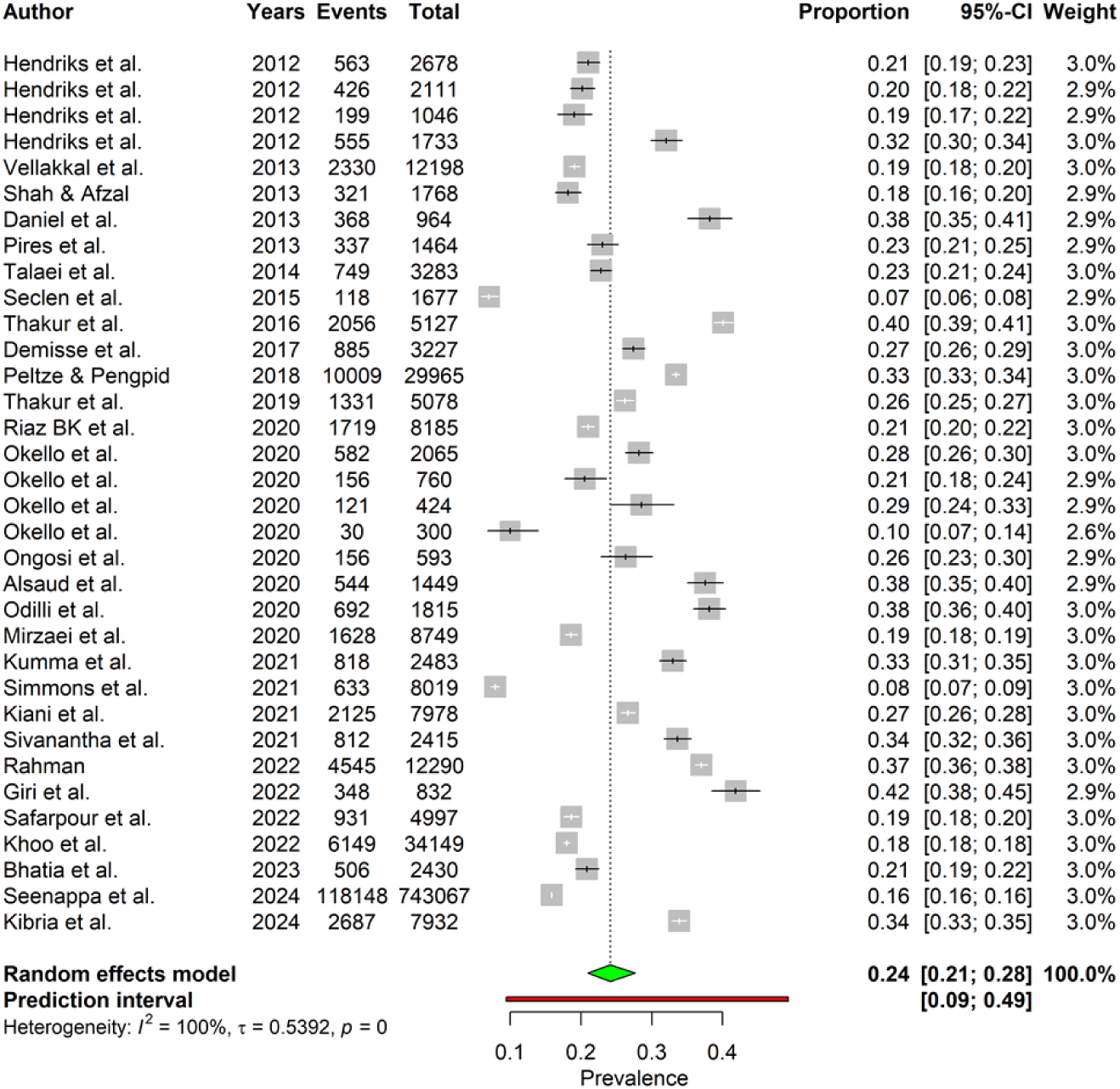
Forest plot illustrating the combined prevalence of hypertension from 34 studies.

In **Figure 4**, it can be observed that 23 studies on the prevalence of DM were included in the global analysis of this meta-analysis. Using the random-effects model, the estimated combined proportion was 0.11 (95% CI: 0.10; 0.13). The prediction for future populations with similar characteristics was estimated to range between 0.05 and 0.23.

**Fig.4.**
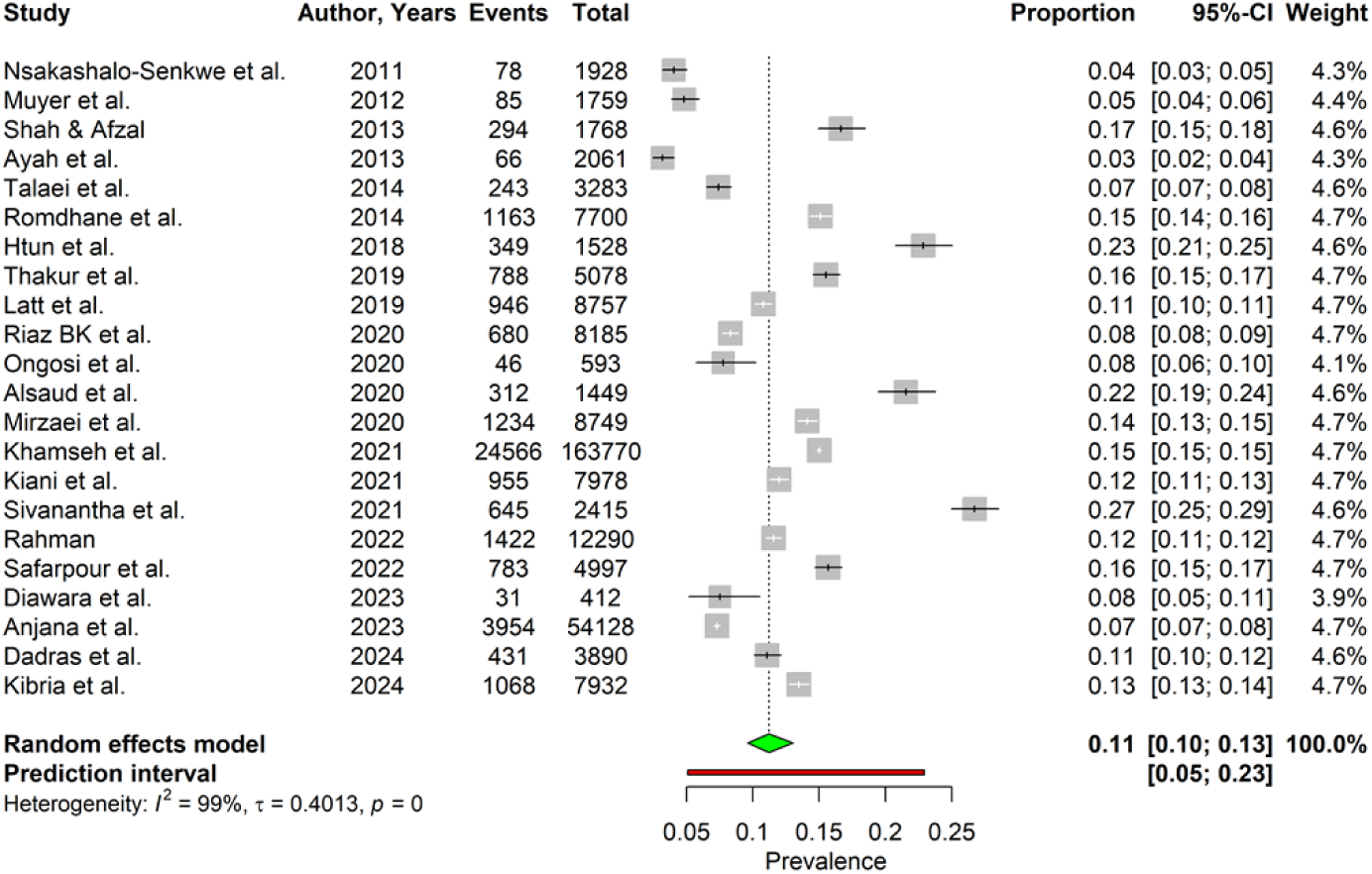
Forest plot illustrating the combined prevalence of DM from 19 studies

### Heterogeneity analysis

Figures 3 **and 4**, which present the forest plot of the combined prevalence of hypertension and DM, respectively, indicated high heterogeneity among the studies. For hypertension, the I² index was 99.8% with a p-value < 0.001 in the Cochrane Q test, while for DM, the I² was 99.4%, also with a p-value < 0.001 in the same test.

### Subgroup and meta-regression analysis

Subgroup analyses were conducted based on study design (cohort and cross-sectional), publication year (2012-2014, 2015-2019, and 2020-2024), sample size, geographic region (Latin America and Caribbean, East Asia and Pacific, Middle East and North Africa, South Asia, and Sub-Saharan Africa), and study quality (Fair, Good, and Poor). The results for the combined prevalence of hypertension and diabetes from the subgroup analysis are shown in **Table 2**.

**Table 2.**
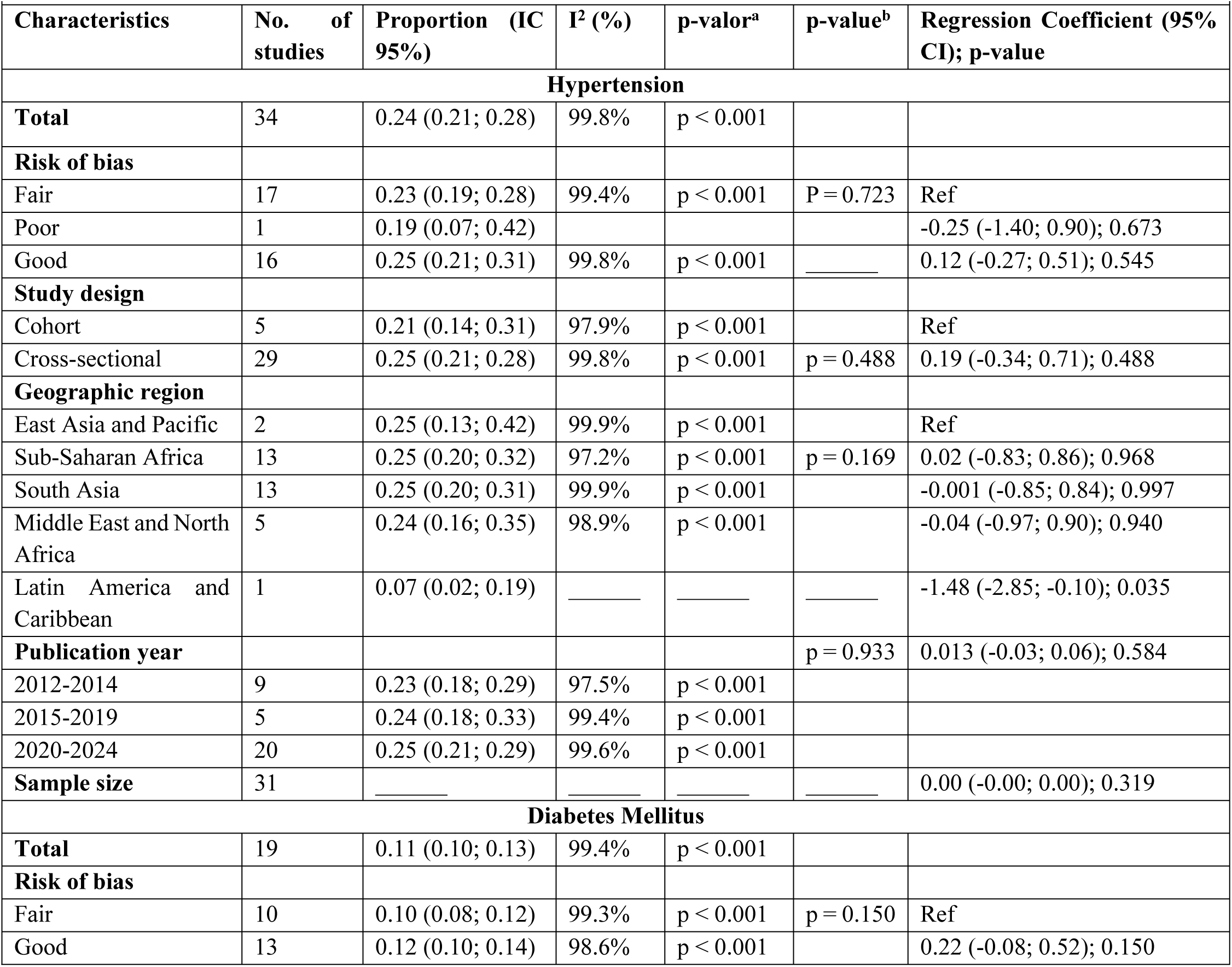
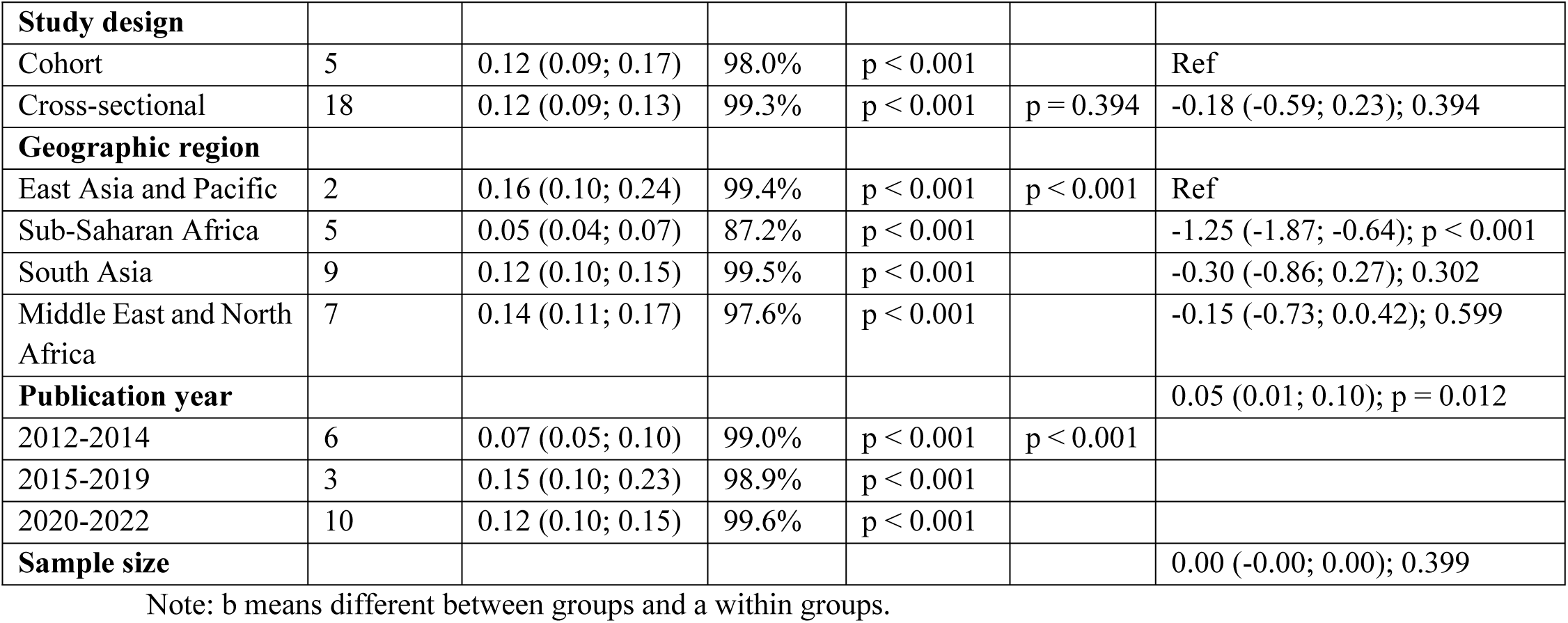
Subgroup analysis and meta-regression for sources of heterogeneity.

In the subgroup analysis by study design, the combined proportion of hypertension was 0.25 (95% CI: 0.21; 0.28) in cross-sectional studies and 0.21 (95% CI: 0.14; 0.31) in cohort studies, both with substantial heterogeneity (I² > 97%). For DM, the combined proportion was 0.12 (95% CI: 0.09; 0.13) in cross-sectional studies and 0.12 (95% CI: 0.09; 0.17) in cohort studies, also with high heterogeneity (I² > 98%). Regarding geographic regions, the combined proportion of hypertension ranged from 0.07 (95% CI: 0.02; 0.19) in Latin America and the Caribbean to 0.25 (95% CI: 0.20; 0.32) in Sub-Saharan Africa. For DM, the East Asia and Pacific region had the highest combined proportion (0.16; 95% CI: 0.10; 0.24), while Sub-Saharan Africa had the lowest (0.05; 95% CI: 0.04; 0.07). When stratified by study quality, Fair studies for hypertension showed a combined proportion of 0.23 (95% CI: 0.19; 0.28), while Good quality studies had a proportion of 0.25 (95% CI: 0.21; 0.31). For DM, Fair studies presented a combined proportion of 0.10 (95% CI: 0.08; 0.12), and Good studies had a proportion of 0.12 (95% CI: 0.10; 0.14)

Univariate meta-regression analysis indicated that for DM, geographic region and publication year were significant sources of heterogeneity. Sub-Saharan Africa displayed significant variability (Coef = -1.25, p < 0.001), and the publication year showed a significant positive association (Coef = 0.05, p = 0.012), suggesting an increasing trend in DM prevalence over time (**Table 2 and Fig.5).**

**Fig. 5.**
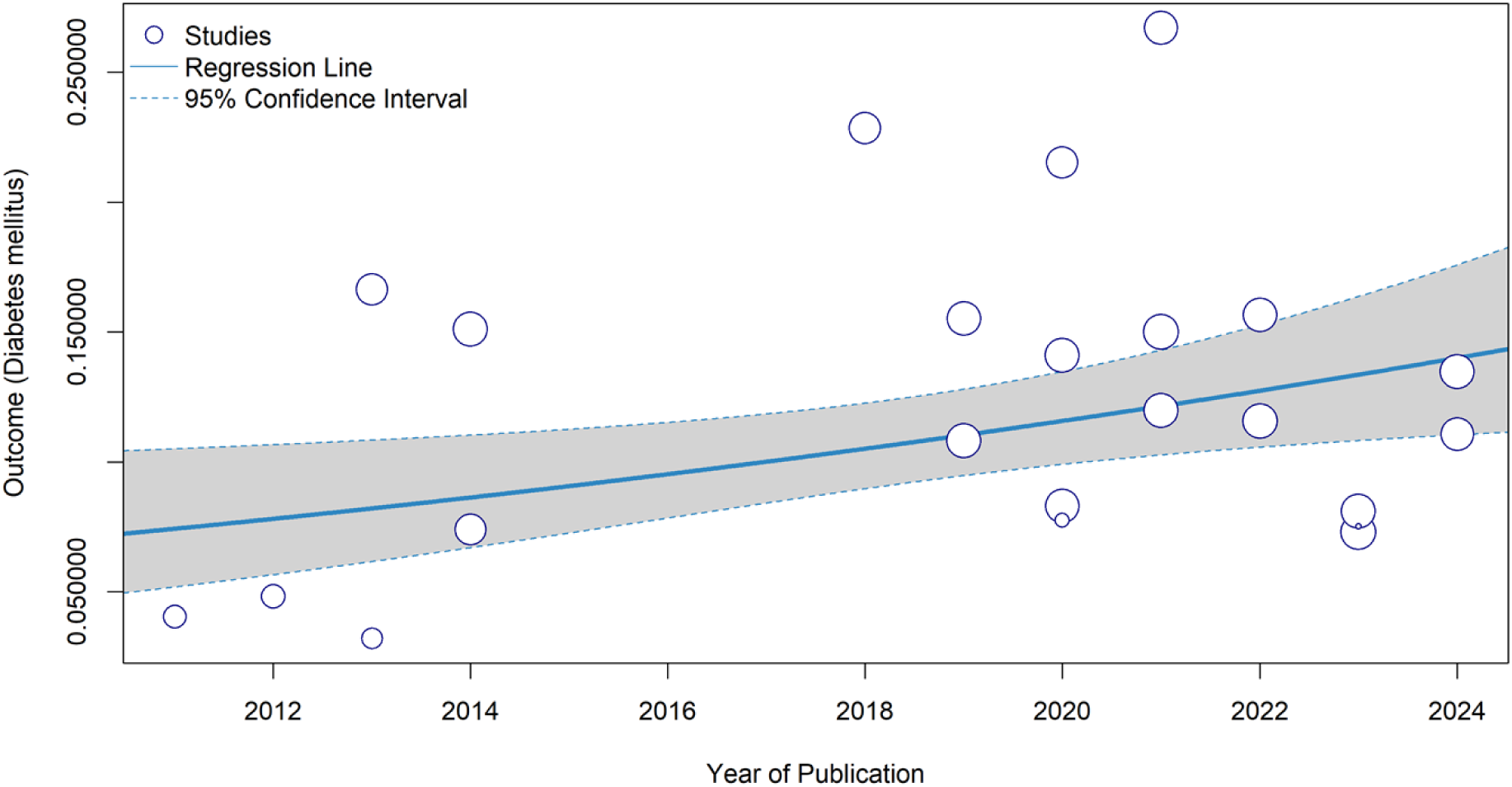
Univariate meta-regression of the prevalence of DM and publication year to analyze heterogeneity among studies and trends over time.

### Publication bias and sensitivity analysis

Publication bias was assessed through the funnel plot and the Egger and Begg regression tests. The funnel plot for hypertension suggested asymmetry in the distribution of studies. The Begg test did not indicate significant bias (z = -1.65, p = 0.099), while the Egger test revealed significant asymmetry (t = 3.66, p = 0.0009), suggesting the possible presence of publication bias. The trim and fill method estimated a combined prevalence of 17.15% (95% CI: 14.67; 19.96) with 14 imputed studies. The uncorrected prevalence was 24.15% (95% CI: 20.97; 27.64). For diabetes, the Egger (t = -1.35, p = 0.190) and Begg (z = -0.50, p = 0.616) tests did not indicate the presence of publication bias, suggesting that the included studies did not show significant asymmetry in the funnel plot. Furthermore, the sensitivity analysis demonstrated that no study significantly influenced the combined results of this meta-analysis (**Fig. S3: Fig.7 and S4 in the supplements**).

### Predictors for chronic non-communicable diseases

#### Risk factors for hypertension

##### Sociodemographic Factors

Sociodemographic factors significantly associated with hypertension include age, male sex, marital status, education level, and economic conditions. The meta-analysis revealed that the risk of hypertension increases considerably with age, with combined ORs of 2.08 (95% CI: 1.87–2.33) for individuals aged 25-34 years, 2.36 (95% CI: 1.59–3.50) for those aged 35-44 years, 3.03 (95% CI: 2.01–4.56) for 45-54 years, and 2.64 (95% CI: 1.62–4.32) for those aged 55 years or older. Age was associated with a 3% increase in hypertension risk for each additional year, as reported by Kumma et al. (2021) with an OR of 1.03 (95% CI: 1.01–1.04) [10]. Men presented a higher risk of hypertension, with a combined OR of 1.56 (95% CI: 1.25–1.95), while women showed no significant association (OR = 0.89; 95% CI: 0.72–1.10). Divorced participants also had an increased risk of hypertension (OR = 1.51; 95% CI: 1.03–2.20).

Economic conditions were also important, with individuals from wealthier classes having a significantly higher risk of hypertension (OR = 1.18; 95% CI: 1.10–1.26), while those with intermediate economic conditions also showed a slightly elevated risk (OR = 1.11; 95% CI: 1.08– 1.15) **(see Table 3)**.

**Table 3.** Risk factors for hypertension: a meta-analysis.

| **Table 3. Risk factors for hypertension: a meta-analysis** |  |  |  |
| --- | --- | --- | --- |
| **Risk factors** | **Article Nr** | **Extracted results: reference number: OR (95% CI) through the log** | **Pooled ORs (IC 95%), p-value, I<sup>2</sup>** |
| **Sociodemographic and Economic** |  |  |  |
| Age (in years) |  |  |  |
| Age | 2 | [10]: 1.04 (1.02; 1.04); |  |
| 25-34 | 2 | [3]: 2.11 (1.88; 2.37); [18]: 1.83 (1.27; 2.64) | 2.08 (1.87; 2.33), p<0.001, 0%. |
| 35-44 | 4 | [3]: 3.35 (2.99; 3.76); [28]: 2.25 (0.92; 5.52); [29]: 1.36 (0.88; 2.09); [18]: 2.56 (1.75; 3.74) | 2.36 (1.59; 3.50), p < 0.001, 82.7% |
| 45-54 | 7 | [3]: 1.46 (1.35; 1.58); [28]: 1.06 (0.32; 1.79); [18]: 1.37 (0.98; 1.75); [4]: 1.81 (1.80; 1.82); [32]: 1.03 (0.91; 1.15); [25]: 1.08 (0.74; 1.58); [24]: 2.35 [2.27; 2.43) | 3.03 (2.014; 4.56), p < 0.001, 99.8% |
| 55 or more | 10 | [3]: 5.21 (4.64; 5.85); [28]: 9.43 (4.05; 21.94); [29]: 3.54 (1.56; 8.02); [30]: 0.90 (0.75; 1.08); [18]: 4.68 (3.16; 6.94); [18]: 5.56 (3.71; 8.34); [5]: 1.22 (1.17; 1.28); [25]: 1.27 (0.92; 1.75); [25]: 1.59 (1.03; 2.45); [25]: 2.36 (0.84; 6.61) | 2.64 (1.62; 4.32) , p < 0.001, 98.6% |
| **Gender** |  |  |  |
| Female | 6 | [3]: 1.13 (1.11; 1.15); [5]: 0.90 (0.86; 0.95); [30]: 1.10 (0.83; 1.46); [32]: 0.60 (0.51; 0.71); [25]: 1.15 (0.72; 1.83); [24]: 0.72 (0.69; 0.75) | 0.89 (0.72; 1.10), p = 0.270, 98.9% |
| Male | 4 | [28]: 1.56 (1.15; 2.11); [10]: 1.40 (1.13; 1.74); [29]: 2.37 (1.48; 3.80); [18]: 1.42 (1.18; 1.71) | 1.56 (1.25; 1.95), p < 0.001, 31.4% |
| Place of residence (urban) | 9 | [3]: 1.01 (0.95; 1.07); [10]: 1.20 (0.78; 1.85); [29]: 1.14 (0.68; 1.91); [30]: 1.10 (0.96; 1.25); [18]: 1.31 (1.10; 1.56); [5]: 1.09 (1.05; 1.14); [32]: 0.90 (0.77; 1.06); [24]: 1.01 (0.98; 1.04) | 1.06 (0.98; 1.15), p = 0.149, 66.2% |
| **Marital status** |  |  |  |
| Single | 4 | [28]: 0.22 ( 0.05; 0.97); [30]: 1.10 (0.75; 1.61); [5]: 0.89 (0.84; 0.94); [32]: 2.10 (1.75; 2.52) | 0.98 (0.43; 2.22), p = 0.960, 96% |
| Married | 3 | [10]: 1.00 (0.71; 1.41); [18]: 1.03 (0.80; 1.33); [24]: 1.19 (1.15; 1.24) | 1.18 (1.14; 1.23), p < 0.001, 6.3% |
| Divorced | 6 | [30]: 0.90 (0.59; 1.37); [18]: 1.87 (1.27; 2.75); [18]: 1.09 (0.68; 1.74); [18]: 1.87 (1.27; 2.75); [5]: 1.06 (0.94; 1.19); [32]: 3.10 (2.32; 4.14) | 1.51 (1.03; 2.20), p = 0.034, 91.1% |
| **Education level** |  |  |  |
| Primary | 4 | [3]: 1.00 (0.93; 1.07); [30]: 1.60 (1.26; 2.03); [5]: 1.04 (0.97; 1.11); [32]: 1.10 (0.92; 1.32); [25]: 1.15 (0.79; 1.67) | 1.13 (0.96; 1.33), p = 0.131, 71.8% |
| Secondary | 4 | [3]: 1.01 (0.93; 1.09); [30]: 1.30 (1.08; 1.57); [5]: 0.96 (0.89; 1.03); [32]: 1.00 (0.82; 1.22) | 1.04 (0.92; 1.19), p = 0.517, 66% |
| Higher or more | 6 | [3]: 1.06 (0.96; 1.17); [10]: 1.00 (0.73; 1.36); [10]: 0.90 (0.69; 1.18); [30]: 1.50 (1.22; 1.84); [5]: 0.97 (0.90; 1.05); [32]: 1.00 (0.82; 1.22); [25]: 0.99 (0.59; 1.66) | 1.06 (0.93; 1.20), p = 0.417, 64.6% |
| **Occupation** |  |  |  |
| Employed | 10 | [3]: 0.92 (0.86; 0.98); [29]: 1.07 (0.60; 1.91); [30]: 1.00 (0.76; 1.32); [30]: 1.00 (0.59; 1.68); [5]: 1.04 (0.91; 1.18); [5]: 0.99 (0.94; 1.05); [32]: 1.00 (0.68; 1.46); [32]: 0.80 (0.62; 1.03); [25]: 0.66 (0.44; 0.99); [24]: 0.92 (0.89; 0.95) | 0.94 (0.91; 0.96), p < 0.001, 29.2% |
| Self-employed | 3 | [29]: 1.72 (1.06; 2.80); [5]: 0.97 (0.91; 1.03); [32]: 1.10 (0.88; 1.37) | 1.14 (0.84; 1.55), p = 0.389, 68% |
| Unemployed | 2 | [25]: 0.78 (0.53; 1.15); [26]: 1.43 (1.19; 1.71) | 1.09 (0.62; 1.92), p = 0.776, 86.8% |
| Others | 2 | [5]: 1.01 (0.95; 1.078); [32]: 1.80 (1.41; 2.29) | 1.33 (0.76; 2.32), p = 0.312, 95% |
| **Economic conditions** |  |  |  |
| Poorer | 2 | [3]: 1.06 (0.97; 1.16); [25]: 0.91 (0.58 1.43) | 1.05 (0.96; 1.15), p = 0.267, 0% |
| Middle | 3 | [3]: 1.11 (1.01; 1.22); [25]: 1.51 (0.98; 2.33); [24]: 1.11 (1.07; 1.15) | 1.11 (1.083; 1.15) p < 0.001, 0% |
| Richer/Richest | 4 | [3]: 1.17 (1.06; 1.29); [3]: 1.18 (1.07; 1.31); [25]: 1.34 (0.86; 2.08); [25]: 1.09 (0.66; 1.81) | 1.18 (1.10; 1.26) p < 0.001, 0% |
| Religion (none) | 1 | [18]: 1.16 (0.95; 1.42) |  |
| Yes | 2 | [25]: 0.83 (0.51; 1.35) |  |
| **Health/Lifestyle-Related Predictors** |  |  |  |
| BMI |  |  |  |
| Obese | 5 | [3]: 2.41 (2.11; 2.75); [10]: 2.50 (1.39; 4.48); [29]: 2.97 (1.68 5.25); [18]: 2.62 (1.70; 4.03); [24]: 1.63 (1.58; 1.68) | 2.19 (1.76; 2.73), P < 0.001, 90.4% |
| Overweight | 3 | [3]: 2.37 (2.14; 2.62); [29]: 1.87 (1.19; 2.93); [18]: 2.29 (1.64; 3.19) | 2.34 (2.13; 2.57), p < 0.001, 0% |
| Normal | 2 | [3]: 1.46 (1.23; 1.73); [18]: 1.51 (1.12; 2.03); | 1.47 (1.27; 1.70), p < 0.001, 0% |
| Overweight/Obese | 2 | [25]: 1.47 (0.94; 2.29); [24]: 1.49 (1.44; 1.54) | 1.49 (1.44; 1.54), p < 0.001, 0% |
| Prediabetes | 1 | [29]: 1.00 (0.52; 1.95) |  |
| Diabetes | 1 | [29]: 2.37 (0.75; 7.50); [25]: 0.84 (0.41; 1.72; [24]: 2.47 (2.31; 2.64) | 1.74 (0.88; 3.46), p = 0.112, 76.6% |
| Stroke | 1 | [25]: 0.13 (0.03; 0.57) |  |
| Waist-to-hip ratio | 1 | [29]: 1.11 (0.73; 1.69) |  |
| Waist circumference |  | [29]: 2.07 (0.75; 5.71) |  |
| Smoking | 6 | [28]: 3.18 (1.67; 6.05); [29]: 1.09 (0.42; 2.82); [25]: 1.88 (1.07; 3.29); [25]: 0.63 (0.22; 1.80); [25]: 1.34 (0.93; 1.94); [24]: 1.01 (0.98; 1.05) | 1.38 (0.90; 2.12), p = 0.145, 75% |
| Occasionally | 1 | [25]: 0.63 (0.22; 1.79) |  |
| Alcohol consumption | 4 | [10]: 2.10 (0.90; 4.90); [29]: 1.47 (0.72; 3.01); [25]: 1.13 (0.71; 1.79); [24]: 1.38 1.33; 1.44) | 1.38 (1.33; 1.44), p < 0.001, 0% |
| Vegetable consumption (daily) | 1 | [10]: 0.6 (0.4; 1.1) |  |
| Total cholesterol (mg/dl and/or mmol/l) | 2 | [10]: 1.20 (1.10; 1.30); [18]: 1.00 (1.00; 1.01) | 1.09 (0.92; 1.30), p = 0.334, 95% |
| Physical activity (Moderate/High) | 3 | [25]: 1.25 (0.91; 1.71); [23]0.53 (0.34; 0.88); [25]: 1.05 (0.73; 1.52) | 0.92 (0.56; 1.50), p = 0.724, 75.1% |
| Sedentary | 1 | [26]: 1.44 (1.15; 1.81) |  |
| Normal | 1 | [11]: 1.00 (0.70; 1.40) |  |
| Blood sugars (mg/dl and/or Mmol (l)) | 2 | [10]: 1.10 (1.01; 1.20); [18]: 1.01 (1.01; 1.02) | 1.04 (0.96; 1.12), p = 0.303, 73% |
| Salt consumption | 1 | [28]: 1.83 (1.03; 3.35) |  |
| **Psychological factors** |  |  |  |
| Stress | 1 | [28]: 1.77 (1.15; 2.37) |  |

##### Health and lifestyle-related factors

Health and lifestyle-related factors significantly associated with hypertension included alcohol consumption, BMI, and sedentary behavior. The risk of hypertension increased progressively with BMI, with ORs of 1.47 (95% CI: 1.27–1.70) for individuals with normal BMI, 2.34 (95% CI: 2.13– 2.57) for overweight, and 2.19 (95% CI: 1.76–2.73) for obesity. Alcohol consumption was also associated with an increased risk of hypertension, with an OR of 1.38 (95% CI: 1.33–1.44). Sedentary behavior increased the risk of hypertension, with an OR of 1.44 (95% CI: 1.15–1.81). Additionally, salt consumption was identified as a risk factor, with an OR of 1.83 (95% CI: 1.03–3.35), and stress was also significantly associated with an increased risk of hypertension, with an OR of 1.77 (95% CI: 1.15–2.37) [28] **(see table 3)**.

#### Risk factors for diabetes mellitus

##### Sociodemographic factors

Sociodemographic factors significantly associated with diabetes mellitus (DM) include age, religion, economic conditions, and marital status. The risk of DM increases with age, with combined odds ratios (OR) of 2.66 (95% CI: 1.93–3.67) for individuals aged 35-44 years, 4.09 (95% CI: 3.08–5.42) for those aged 45-54 years, and 4.04 (95% CI: 2.47–6.60) for those aged 55 years or older. Age was associated with a 4% increase in DM risk for each additional year (OR = 1.04; 95% CI: 1.01–1.06). Individuals with no religious affiliation had an elevated risk of DM, with a combined OR of 1.90 (95% CI: 1.90–1.90). Marital status was also statistically significant, with divorced individuals having a higher risk of DM (OR = 1.79; 95% CI: 1.04–3.09). In terms of economic conditions, individuals in the higher-income group (richer/richest) were at greater risk for DM, with a pooled OR of 1.34 (95% CI: 0.94–1.91). Additionally, the risk was higher for urban residents (OR = 1.16; 95% CI: 1.00–1.35) compared to rural residents (see Table 4).

**Table 4.** Risk factors for diabetes mellitus: a meta-analysis.

| **Table 4.** Risk factors for diabetes mellitus: a meta-analysis |  |  |  |
| --- | --- | --- | --- |
| **Predictores** | **Article Nr** | **Extracted results: reference number: OR (95% CI) through the log** | **Pooled ORs (IC 95%), p-value, I<sup>2</sup>** |
| **Sociodemographic and economic** |  |  |  |
| Age (in years) | 1 | [31]: 1.05 (1.04; 1.08); [23]: 1.02 (1.00; 1.04) | 1.04 (1.01; 1.06), p = 0.013, 77.1% |
| 35-44 | 4 | [3]: 2.91 (2.39; 3.55); [4]: 3.30 (3.25; 3.35); [29]: 1.51 (0.86; 2.65); [22]: 2.49 (1.26; 4.92) | 2.66 (1.93; 3.67), p < 0.001, 69% |
| 45-54 | 7 | [3]: 3.96 (3.24; 4.84); [27]: 5.22 (1.09; 25.01); [11]: 2.71 (2.56; 2.86); [4]: 7.70 (7.65; 7.75); [29]: 3.59 (1.78; 7.25); [22]: 3.80 (2.00; 7.23); [32]: 3.50 (2.82; 4.34) | 4.09 (3.08; 5.42), p < 0.001, 100% |
| 55 or more | 7 | [3]: 3.96 (3.19; 4.92); [3]: 3.85 (3.01; 4.93); [27]: 4.44 (0.84 23.59); [11]: 4.86 (4.57; 5.12); [11]: | 4.04 (2.47; 6.60), p < 0.001, 100% |
|  |  | 5.73 (5.03; 6.52); [5]: 1.13 (1.06; 1.20); [4]: 8.70 (8.65; 8.75) |  |
| Gender (Female) | 4 | [11]: 1.10 (1.00; 1.21); [5]: 0.90 (0.84; 0.96); [3]: 0.95 (0.83; 1.09); [32]: 0.90 (0.64; 1.27) | 0.97 (0.88; 1.074), p = 0.574, 74.1% |
| Male | 1 | [31]: 1.42 (0.89; 2.27) |  |
| Place of residence (urban) | 5 | [3]: 0.99 (0.88; 1.12); [30]: 1.40 (1.18; 1.67); [5]: 1.03 (0.97; 1.09); [32]: 1.20 (1.01; 1.42); [23]: 1.49 (1.00; 2.21) | 1.16 (1.00; 1.35), p = 0.054, 76.3% |
| Rural | 2 | [11]: 0.79 (0.73; 0.85); [25]: 1.06 (0.73; 1.54) | 0.86 (0.66; 1.11), p = 0.253, 55.6% |
| **Marital status** |  |  |  |
| Single | 3 | [30]: 1.00 (0.63; 1.58); [5]: 0.85 (0.80; 0.90); [32]: 3.00 (1.85; 4.87) | 1.33 (0.63; 2.85), p = 0.455, 92% |
| Married | 2 | [11]: 1.02 (0.97; 1.08); [23]: 0.97 (0.49; 1.91) | 1.02 (0.97; 1.06), p = 0.476, 0% |
| Divorced | 5 | [30]: 0.90 (0.50; 1.61); [5]: 1.06 (0.97; 1.16); [4]: 2.90 (2.85; 2.95); [32]: 3.70 (2.09; 6.56); [23]: 1.83 (0.77; 4.35) | 1.79 (1.04; 3.09), p = 0.034, 99.2% |
| **Education level** |  |  |  |
| Primary | 7 | [3]: 1.25 (1.08; 1.45); [30]: 1.40 (1.02; 1.93); [31]: 0.71 (0.43; 1.18); [11]: 0.87 (0.82; 0.93); [11]: 0.84 (0.78; 0.90); [5]: 1.04 (0.95; 1.13); [32]: 1.50 (1.14; 1.97) | 1.06 (0.87; 1.28), p = 0.594, 89% |
| Secondary | 6 | [3]: 1.21 (1.02; 1.44); [30]: 0.90 (0.72; 1.13); [31]: 0.69 (0.39; 1.22); [11]: 0.77 (0.73; 0.81); [5]: 1.01 (0.91; 1.12); [4]: 1.20 (0.82; 1.75) | 0.96 (0.80; 1.14), p = 0.614, 89% |
| Higher or more | 6 | [3]: 1.13 (0.92; 1.38); [29]: 0.98 (0.58; 1.66); [30]: 1.10 (0.88; 1.37); [11]: 0.71 (0.68; 0.75); [5]: 0.98 (0.88; 1.09); [32]: 1.50 (1.14; 1.97) | 1.02 (0.83; 1.25), p = 0.831, 93% |
| **Occupation** |  |  |  |
| Employed | 7 | [3]: 0.83 (0.72; 0.95); [29]: 0.43 (0.20; 0.92); [30]: 1.00 (0.68; 1.46); [30]: 1.50 (0.70; 3.21); [5]: 0.99 (0.92; 1.07); [32]: 0.70 (0.42; 1.16); [32]: 0.50 (0.33; 0.76) | 0.80 (0.59; 1.08), p = 0.141, 71% |
| Self-employed | 4 | [29]: 0.57 (0.33; 0.99); [30]: 1.20 (0.87; 1.65); [32]: 0.60 (0.40; 0.90) | 0.83 (0.59; 1.17), p = 0.286, 71% |
| Unemployed | 1 | [26]: 1.82 (1.55; 2.13) |  |
| Others | 4 | [30]: 0.90 (0.64; 1.27); [5]: 1.12 (1.02; 1.23); [4]: 2.90 (2.85; 2.95); [32]: 0.70 (0.47; 1.04) | 1.22 (0.66; 2.25), p = 0.524, 99% |
| **Economic conditions** |  |  |  |
| Poorer | 3 | [3]: 0.97 (0.77; 1.22); [31]: 1.70 (1.08; 2.67); [11]: 1.06 (0.99; 1.14) | 1.14 (0.84; 1.55) p = 0.395, 57.9% |
| Middle | 3 | [3]: 1.22 (0.98; 1.52); [31]: 1.30 (0.84; 2.00); [11]: 1.06 (0.98; 1.15) | 1.08 (1.01; 1.17), p = 0.034, 4.6% |
| Richer/Richest | 4 | [3]: 1.53 (1.23; 1.90); [3]: 2.15 (1.73; 2.68); [11]: 1.00 (0.90; 1.11); [11]: 1.02 (0.94; 1.10) | 1.34 (0.94; 1.91), p = 0.107, 94.2% |
| Religion (none) | 1 | [4]: 1.90 (1.90; 1.90) |  |
| **Health/Lifestyle-Related Predictors** |  |  |  |
| BMI |  |  |  |
| + BMI | 1 | [31]: 1.20 (1.15; 1.25) |  |
| Obese | 5 | [11]: 1.83 (1.70; 1.97); [3]: 2.22 (1.71; 2.89); [5]: 3.01 (1.67; 5.42); [31]: 1.20 (1.15; 1.25); [22]: 1.76 (0.64; 4.86); [23]: 2.54 (1.37; 4.70) | 1.88 (1.43; 2.46), p < 0.001, 95.9% |
| Overweight | 5 | [11]: 1.43 (1.34; 1.53); [3]: 2.00 (1.61; 2.48); [29]: 1.10 (0.59; 2.05); [22]: 0.82 (0.30; 2.25); [23]: 2.12 (1.10; 4.09) | 1.54 (1.11; 2.12), p = 0.008, 66.5% |
| Normal | 2 | [3]: 1.41 (1.16; 1.71); [22]: 0.51 (0.20; 1.33) | 0.96 (0.38; 2.42), p = 0.928, 76% |
| Underweight | 2 | [11]: 0.39 (0.26; 0.58); [23]: 1.24 (0.26; 5.90) | 0.42 (0.29; 0.61), p < 0.001, 49.7% |
| Family history of Diabetes | 1 | [27]: 14.13 (6.10; 32.75) |  |
| Hemoglobin concentration | 1 | [31]: 1.02 (1.01; 1.03) |  |
| Hypertension | 6 | [27]: 2.06 (0.86; 4.94); [29]: 1.52 (0.78; 2.95); [22]: 2.57 (1.44; 4.58); [22]: 0.98 (0.34; 2.83); [22]: 1.84 (0.75; 4.50); [23]: 1.82 (1.16; 2.86) | 1.85 (1.41; 2.43), p < 0.001, 0% |
| Prehypertension | 1 | [29]: 1.06 (0.57; 1.98) |  |
| Waist-to-hip ratio | 2 | [29]: 2.52 (1.43; 4.44); [11]: 2.20 (2.03; 2.39) | 2.21 (2.03; 2.39), p < 0.001, 0% |
| Smoking (yes) | 1 | [31]: 1.98 (1.22; 2.92) |  |
| Smoking (former) | 1 | [11]: 1.0 (0.87; 1.16) |  |
| Smoking (current) | 1 | [11]: 0.82 (0.72; 0.92) |  |
| Alcohol consumption | 1 | [31]: 0.48 (0.33; 1.03) |  |
| Total cholesterol (mg/dl and/or mmol/l) | 2 | [10]: 1.20 (1.103; 1.30); [18]: 1.00 (1.00; 1.01) | 1.09 (0.92; 1.30), p = 0.334, 95% |
| Low-density lipoprotein (mg/dl) (High ($\geq 130$)) | 2 | [29]: 1.11 (0.57; 2.15); [22]: 1.23 (0.67; 2.26) | 1.17 (0.75; 1.83), p = 0.483, 0% |
| Physical activity (Moderate/High) | 2 | [31] : 1.19 (0.76; 1.87); [11]: 0.64 (0.60; 0.68); [11]: 0.79 (0.74; 0.84) | 0.80 (0.58; 1.10), p = 0.169, 92.5% |
| Sedentary | 1 | [26]: 2.23 (1.87; 2.87) |  |
| **Psychological factors** |  |  |  |
| Opium consumption | 1 | [11]: 1.0 (0.92; 1.08) |  |
| **Simple infection** |  |  |  |
| Taenia spp. | 1 | [31]: 2.98 (1.10; 8.05) |  |
| Strongyloides stercoralis | 1 | [31]: 0.65 (0.15; 2.72) |  |
| Minute intestinal flukes | 1 | [31]: 1.20: (0.57; 2.52) |  |
| Opisthorchis viverrine | 1 | [31]: 0.87 (0.58; 1.28) |  |
| Hookworm | 1 | [31]: 0.94 (0.42; 2.10) |  |

##### Healthy lifestyle factors

Among health and lifestyle-related predictors, BMI, family history of DM, hypertension, hemoglobin concentration, waist-to-hip ratio, smoking, physical activity, and simple infections were highlighted. The risk of DM increased progressively with BMI, with ORs of 1.54 (95% CI: 1.11–2.12) for overweight and 1.88 (95% CI: 1.43–2.46) for obesity. Individuals with a family history of DM had a significantly higher risk, with an OR of 14.13 (95% CI: 6.10–32.75). Hemoglobin concentration was also identified as a relevant factor, with a 2% increase in risk for each additional unit of hemoglobin (OR: 1.02; 95% CI: 1.01–1.03). Smoking was associated with an increased risk of DM, with an OR of 1.98 (95% CI: 1.22–2.92). Individuals with hypertension had an increased risk of DM (OR: 1.85; 95% CI: 1.41–2.43), as did those with a high waist-to-hip ratio (OR: 2.21; 95% CI: 2.03–2.39

Smoking was also associated with an increased risk of DM, with an OR of 1.98 (95% CI: 1.22–2.92), while simple infections, such as Taenia spp., increased the risk of DM by 2.98 times (95% CI: 1.10– 8.05). On the other hand, sedentary individuals had a higher risk of DM (OR = 2.23; 95% CI: 1.87– 2.87) (see Table 4).

## Discussion

This meta-analysis investigated the prevalence of NCDs, such as hypertension and DM, as well as their associated risk factors in countries classified as LMIC by the World Bank. The estimated prevalence of hypertension was 24% (95% CI: 21% - 28%), with a prediction of up to 49% for similar populations. For DM, the estimated prevalence was 11% (95% CI: 10% - 13%), with a prediction of up to 23% in similar populations. The prevalence of hypertension found in this study was consistent with estimates from other primary studies. In a study conducted in 44 LMICs, the prevalence was 17.5% [33]. Another study conducted in 92 countries reported a prevalence of 34% [34]. Regarding DM, the prevalence was similar to that found in other studies, with 8.7% in one study in some LMICs [35], 9.0% in 55 LMICs [36], and 7.5% in 29 LMICs [33]. When analyzing regional prevalences of hypertension in this meta-analysis, no statistically significant differences were found between regions: Sub-Saharan Africa (25%), East Asia and Pacific (25%), South Asia (26%), Middle East and North Africa (24%), although Latin America and the Caribbean had a lower proportion (7.0%). These results are comparable to those of other studies, where the prevalence in the South Asia region reached 29.3%, in Sub-Saharan Africa 27.9%, in North Africa and the Middle East 30.6%, and in East Asia 30.6% [37], with up to 33.8% for South Asia, 24.8% for Sub-Saharan Africa, 26.3% for North Africa and the Middle East, and 30.7% for East Asia [38]. Regarding DM, our results were similar to those found in a primary study, which reported a prevalence of 5.2% in the Sub-Saharan Africa region. However, differences were observed in other regions, such as Southeast Asia and the Pacific, where the prevalence was 7.7% [39], with high prevalences in South Asia, reaching 19.0% [40, 41], and a prevalence of 12.2% in the Middle East and North Africa, with a prediction of up to 96% increase [42], and 9.8 for Latin America and the Caribbean, specifically Peru [40]. NCDs, such as hypertension and DM, represent a rapidly growing health problem in LMICs, where the rate of prevalence increase in the last two decades has surpassed that of high-income countries, accompanying the economic growth of these nations [39, 43]. These findings highlight a significant public health concern, underscoring the urgency of more effective preventive interventions and management strategies to reduce the risk and complications associated with these diseases, such as heart attack and stroke.

Several risk factors associated with hypertension and DM were identified in this study. For hypertension, the factors include age, sex, marital status, economic conditions, BMI, alcohol consumption, salt intake, and stress. For DM, the risk factors found were age, economic conditions, physical activity, BMI, family history of diabetes, hypertension, hemoglobin concentration, waist-to-hip ratio, smoking, and simple infections. Regarding age, the meta-analysis identified an increased risk of hypertension and DM with increasing age. These results were similar to those found in previously published studies [44–47]. Therefore, it is advisable to design a health education and promotion mechanism to improve disease controls as patients age. The risk of hypertension was higher in adult males. Similar evidence was found in previous studies [44, 48, 49]. This association may be explained by behavioral differences regarding healthcare between these groups, as women tend to seek healthcare more often than men. Furthermore, the difference can also be justified by the fact that men are more likely to engage in unhealthy lifestyle practices such as smoking, alcohol consumption, and poor eating habits [44, 50].

In this meta-analysis, marital status showed a significant association with hypertension and diabetes mellitus, with married and divorced individuals having a higher risk for these diseases, consistent with previous studies[51, 52]. These associations can be explained by the physiological and psychological stress linked to marital responsibilities and economic pressures, which increase the risk of hypertension in married individuals. Moreover, marital breakdown can lead to unhealthy behaviors, such as smoking, alcohol consumption, and poor diet, negatively affecting mental health and increasing the risk of hypertension and diabetes, possibly due to inflammatory processes and altered glycemic regulation [51, 52]. In this review, alcohol consumption emerged as a risk factor for hypertension, while smoking was identified as a risk factor for diabetes. These behaviors, often associated with marital breakdown, may exacerbate physiological stress responses, further contributing to the elevated risk of these chronic conditions.

The risk of hypertension increased with higher BMI, consistent with findings from previous studies [45, 53–55]. Scientific evidence suggests that inflammatory processes play an important role in the development of hypertension, possibly explaining the association between high BMI and hypertension. Adipose cells, by producing inflammatory cytokines, contribute to increased blood pressure and target organ damage. Increased adipose tissue can also reduce the production of nitric oxide, crucial for vascular tone control, whose decrease is associated with endothelial dysfunction and hypertension [56, 57]. This research also found an increased risk of DM with higher BMI. Consistent findings were found in other previously conducted studies [46, 58, 59]. It is scientifically established that DM develops from insulin deficiency and/or insulin resistance. Some studies have shown that increased BMI induces chronic inflammation [60]. Insulin resistance is partly a result of adipocyte dysregulation, which releases cytokines such as TNF-α and adiponectin [61]. While TNF-α promotes insulin resistance, adiponectin improves the body’s insulin sensitivity. The accumulation of visceral fat, common with weight gain, increases the production of TNF-α and decreases that of adiponectin, leading to insulin resistance and increased blood glucose levels. These mechanisms are fundamental in metabolic syndrome, characterized by visceral obesity, hyperglycemia, dyslipidemia, and hypertension, which are interrelated [62]. In our study, waist-to-hip ratio, hemoglobin concentration, hypertension, and smoking were risk factors associated with DM. A high waist-to-hip ratio reflects abdominal fat accumulation, which is strongly linked to insulin resistance. Hemoglobin concentration can indicate chronic glycemic control, as this condition is related to chronic inflammation and oxidative stress related to diabetes. Hypertension and smoking, in turn, contribute to systemic inflammation and endothelial dysfunction, both factors that increase the risk of developing DM [63, 64].

In this study, an increased risk of hypertension was observed with added salt intake and stress. These findings were consistent with findings in previously conducted studies [45, 65]. Excessive salt intake is associated with the development of hypertension due to various physiological mechanisms, such as water retention by the kidneys, increasing blood volume, and consequently raising blood pressure [66]. Additionally, excess sodium causes vasoconstriction, impairs endothelial function, promotes oxidative stress and arterial inflammation, and activates the renin-angiotensin-aldosterone system, all contributing to increased blood pressure [67]. The association of stress with hypertension can be explained by the increase in vasoconstrictor hormones that raise blood pressure [68].

Based on the results of the meta-analysis, it is evident that without urgent interventions, LMICs will face a continuous increase in NCDs, such as hypertension and DM, where the rates of awareness, diagnosis, treatment, and control are worryingly low. The limitations in the healthcare systems of these countries are contributing to a substantial excess mortality associated with these conditions. Lifestyle-related risk factors, such as obesity, high sodium intake, alcohol consumption, physical inactivity, and inadequate diet, have reached alarming levels [69, 70]. Therefore, it is crucial to implement national prevention, early detection, and treatment programs to reverse the current epidemic trends. Additionally, intervention programs are needed to address health disparities, ensuring that the most vulnerable populations receive the necessary support to mitigate risks and improve health outcomes.

## Limitations

There are some limitations to be considered in this review. First, only observational studies were included, excluding case-control studies, and only studies with estimates adjusted for potential confounding factors were considered in our meta-analysis. It was not possible to perform meta-analyses for several less-reported factors, which were identified by only one study. The meta-analyses conducted for other factors provided combined effect estimates, offering information on the strength of associations and heterogeneity between studies. Additionally, there was significant heterogeneity among the included studies, due to the diversity of studied populations, such as geographic region and publication year. Another limitation was the evidence of publication bias, as indicated by Egger’s test. Although the trim and fill method imputed 14 additional studies and estimated a lower prevalence of hypertension (17.15%, 95% CI: 14.67; 19.96), we opted to consider the unadjusted results (24.15%, 95% CI: 20.97; 27.64). This decision was made because the studies imputed by the trim and fill are estimates based on assumptions, which could compromise the robustness of the findings. Despite these limitations, most of the included studies were of good quality (55.3%), with only 2.6% being of poor quality. The results of this meta-analysis may be useful for formulating effective health policies, such as prevention and early detection programs, aimed at improving the quality of life of individuals living in low- and middle-income countries and helping to reverse current NCDs trends.

## Conclusion

NCDs, such as hypertension and DM, represent a growing public health challenge in LMICs. Identifying associated risk factors is crucial for the development of effective prevention and control strategies. Our findings may assist policymakers in identifying high-risk groups and recommending appropriate prevention strategies.

## Data Availability

The data underlying the findings of this study are fully available and can be accessed without restriction. All relevant data are included within the manuscript and its supporting information files. Additional datasets generated or analyzed during the current study are available from the corresponding author upon reasonable request.

## Supporting information

**S1.** PRISMA Checklist

**S2.** NIH Quality Assessment Tool for Observational Cohort and Cross-Sectional Studies

**S3.** Sensitivity analysis regarding hypertension and DM

**S4.** Forest plot evaluating the risks of publication bias in the articles

## Author Contributions

**Conceptualization:** Sancho Pedro Xavier, Ana Raquel Gotine, Melsequisete Daniel Vasco, Audêncio Victor

**Data curation:** Sancho Pedro Xavier, Ana Raquel Gotine, Melsequisete Daniel Vasco, Audêncio Victor

**Formal analysis:** Sancho Pedro Xavier

**Methodology:** Sancho Pedro Xavier, Ana Raquel Gotine, Audêncio Victor

**Project administration:** Sancho Pedro Xavier

**Resources:** Sancho Pedro Xavier

**Supervision:** Sancho Pedro Xavier

**Validation:** Audêncio Victor, Ana Raquel Gotine, Sancho Pedro Xavier

**Writing – original draft**: Sancho Pedro Xavier, Ana Raquel Gotine, Melsequisete Daniel Vasco

**Writing – review & editing**: Sancho Pedro Xavier, Ana Raquel Gotine, Melsequisete Daniel Vasco, Audêncio Victor

## References

1. Sadeghi M, Talaei M, Oveisgharan S, Rabiei K, Dianatkhah M, Bahonar A, Sarrafzadegan N (2014) The cumulative incidence of conventional risk factors of cardiovascular disease and their population attributable risk in an Iranian population: The Isfahan Cohort Study. Adv Biomed Res 3:242

2. Ndubuisi NE (2021) Noncommunicable Diseases Prevention In Low- and Middle-Income Countries: An Overview of Health in All Policies (HiAP). Inquiry 58:46958020927885

3. Rahman MA (2022) Socioeconomic inequalities in the risk factors of noncommunicable diseases (hypertension and diabetes) among Bangladeshi population: Evidence based on population level data analysis. PLoS One 17:e0274978

4. Riaz BK, Islam MZ, Islam ANMS, Zaman MM, Hossain MA, Rahman MM, Khanam F, Amin KMB, Noor IN (2020) Risk factors for non-communicable diseases in Bangladesh: findings of the population-based cross-sectional national survey 2018. BMJ Open 10:e041334

5. Sivanantham P, Sahoo J, Lakshminarayanan S, Bobby Z, Kar SS (2021) Profile of risk factors for Non-Communicable Diseases (NCDs) in a highly urbanized district of India: Findings from Puducherry district-wide STEPS Survey, 2019–20. PLoS One 16:e0245254

6. Alwan A, MacLean DR (2009) A review of non-communicable disease in low-and middle-income countries. Int Health 1:3–9

7. Tawfik GM, Dila KAS, Mohamed MYF, Tam DNH, Kien ND, Ahmed AM, Huy NT (2019) A step by step guide for conducting a systematic review and meta-analysis with simulation data. Trop Med Health 47:46

8. Shamseer L, Moher D, Clarke M, Ghersi D, Liberati A, Petticrew M, Shekelle P, Stewart LA (2015) Preferred reporting items for systematic review and meta-analysis protocols (PRISMA-P) 2015: elaboration and explanation. BMJ 350:g7647

9. Van der Mierden S, Tsaioun K, Bleich A, Leenaars CHC (2019) Software tools for literature screening in systematic reviews in biomedical research. ALTEX 36:508–517

10. Kumma WP, Lindtjørn B, Loha E (2021) Prevalence of hypertension, and related factors among adults in Wolaita, southern Ethiopia: a community-based cross-sectional study. PLoS One 16:e0260403

11. Khamseh ME, Sepanlou SG, Hashemi-Madani N, et al (2021) Nationwide Prevalence of Diabetes and Prediabetes and Associated Risk Factors Among Iranian Adults: Analysis of Data from PERSIAN Cohort Study. Diabetes Ther Res Treat Educ diabetes Relat Disord 12:2921–2938

12. Higgins JPT, Thompson SG, Deeks JJ, Altman DG (2003) Measuring inconsistency in meta-analyses. BMJ 327:557–560

13. Cochran WG (1954) The combination of estimates from different experiments. Biometrics 10:101–129

14. Higgins JPT, Thompson SG (2002) Quantifying heterogeneity in a meta-analysis. Stat Med 21:1539–1558

15. Egger M, Davey Smith G, Schneider M, Minder C (1997) Bias in meta-analysis detected by a simple, graphical test. BMJ 315:629–634

16. Begg CB, Mazumdar M (1994) Operating characteristics of a rank correlation test for publication bias. Biometrics 50:1088–1101

17. Cooper H, Hedges L V, Valentine JC (2019) The handbook of research synthesis and meta-analysis. Russell Sage Foundation

18. Demisse AG, Greffie ES, Abebe SM, Bulti AB, Alemu S, Abebe B, Mesfin N (2017) High burden of hypertension across the age groups among residents of Gondar city in Ethiopia: a population based cross sectional study. BMC Public Health 17:647

19. Ben Romdhane H, Ali S Ben, Aissi W, Traissac P, Aounallah-Skhiri H, Bougatef S, Maire B, Delpeuch F, Achour N (2014) Prevalence of diabetes in Northern African countries: the case of Tunisia. BMC Public Health 14:86

20. Ayah R, Joshi MD, Wanjiru R, Njau EK, Otieno CF, Njeru EK, Mutai KK (2013) A population-based survey of prevalence of diabetes and correlates in an urban slum community in Nairobi, Kenya. BMC Public Health 13:371

21. Pires JE, Sebastião Y V, Langa AJ, Nery S V (2013) Hypertension in Northern Angola: prevalence, associated factors, awareness, treatment and control. BMC Public Health 13:90

22. Nsakashalo-Senkwe M, Siziya S, Goma FM, Songolo P, Mukonka V, Babaniyi O (2011) Combined prevalence of impaired glucose level or diabetes and its correlates in Lusaka urban district, Zambia: a population based survey. Int Arch Med 4:2

23. Dadras O, Nyaboke Ongosi A, Wang C-W (2024) Prevalence and correlates of diabetes and impaired fasting glucose among adults in Afghanistan: Insights from a national survey. SAGE open Med 12:20503121241238148

24. Seenappa K, Kulothungan V, Mohan R, Mathur P (2024) District-Wise Heterogeneity in Blood Pressure Measurements, Prehypertension, Raised Blood Pressure, and Their Determinants Among Indians: National Family Health Survey-5. Int J Public Health 69:1606766

25. Bhatia M, Dixit P, Kumar M, Dwivedi LK (2023) A longitudinal study of incident hypertension and its determinants in Indian adults aged 45 years and older: evidence from nationally representative WHO-SAGE study (2007-2015). Front Cardiovasc Med 10:1265371

26. Kibria GM Al, Hossen S, Gibson D (2024) The burden of hypertension, diabetes, and overweight/obesity by sedentary work pattern in Bangladesh: Analysis of Demographic and Health Survey 2017-18. PLOS Glob public Heal 4:e0002788

27. Diawara A, Coulibaly DM, Hussain TYA, Cisse C, Li J, Wele M, Diakite M, Traore K, Doumbia SO, Shaffer JG (2023) Type 2 diabetes prevalence, awareness, and risk factors in rural Mali: a cross-sectional study. Sci Rep 13:3718

28. Giri PP, Mohapatra B, Kar K (2022) Prevalence of hypertension and the associated factors among Sabar and Munda tribes of Eastern India. J Fam Med Prim care 11:5065–5071

29. Ongosi AN, Wilunda C, Musumari PM, Techasrivichien T, Wang C-W, Ono-Kihara M, Serrem C, Kihara M, Nakayama T (2020) Prevalence and Risk Factors of Elevated Blood Pressure and Elevated Blood Glucose among Residents of Kajiado County, Kenya: A Population-Based Cross-Sectional Survey. Int J Environ Res Public Health. 10.3390/ijerph17196957

30. Thakur JS, Jeet G, Nangia R, et al (2019) Non-communicable diseases risk factors and their determinants: A cross-sectional statewide STEPS survey, Haryana, North India. PLoS One 14:e0208872

31. Htun NSN, Odermatt P, Paboriboune P, et al (2018) Association between helminth infections and diabetes mellitus in adults from the Lao People’s Democratic Republic: a cross-sectional study. Infect Dis poverty 7:105

32. Thakur JS, Jeet G, Pal A, et al (2016) Profile of Risk Factors for Non-Communicable Diseases in Punjab, Northern India: Results of a State-Wide STEPS Survey. PLoS One 11:e0157705

33. Geldsetzer P, Manne-Goehler J, Marcus M-E, Ebert C, Zhumadilov Z, Wesseh CS, Tsabedze L, Supiyev A, Sturua L, Bahendeka SK (2019) The state of hypertension care in 44 low-income and middle-income countries: a cross-sectional study of nationally representative individual-level data from 1· 1 million adults. Lancet 394:652–662

34. Schutte AE, Srinivasapura Venkateshmurthy N, Mohan S, Prabhakaran D (2021) Hypertension in low-and middle-income countries. Circ Res 128:808–826

35. Dagenais GR, Gerstein HC, Zhang X, McQueen M, Lear S, Lopez-Jaramillo P, Mohan V, Mony P, Gupta R, Kutty VR (2016) Variations in diabetes prevalence in low-, middle-, and high-income countries: results from the prospective urban and rural epidemiological study. Diabetes Care 39:780–787

36. Flood D, Seiglie JA, Dunn M, Tschida S, Theilmann M, Marcus ME, Brian G, Norov B, Mayige MT, Gurung MS (2021) The state of diabetes treatment coverage in 55 low-income and middle-income countries: a cross-sectional study of nationally representative, individual-level data in 680 102 adults. Lancet Heal Longev 2:e340–e351

37. Beaney T, Schutte AE, Stergiou GS, Borghi C, Burger D, Charchar F, Cro S, Diaz A, Damasceno A, Espeche W (2020) May Measurement Month 2019: the global blood pressure screening campaign of the International Society of Hypertension. Hypertension 76:333–341

38. Beaney T, Burrell LM, Castillo RR, Charchar FJ, Cro S, Damasceno A, Kruger R, Nilsson PM, Prabhakaran D, Ramirez AJ (2019) May Measurement Month 2018: a pragmatic global screening campaign to raise awareness of blood pressure by the International Society of Hypertension. Eur Heart J 40:2006–2017

39. Seiglie JA, Marcus M-E, Ebert C, Prodromidis N, Geldsetzer P, Theilmann M, Agoudavi K, Andall-Brereton G, Aryal KK, Bicaba BW (2020) Diabetes prevalence and its relationship with education, wealth, and BMI in 29 low-and middle-income countries. Diabetes Care 43:767–775

40. Shen J, Kondal D, Rubinstein A, et al (2016) A Multiethnic Study of Pre-Diabetes and Diabetes in LMIC. Glob Heart 11:61–70

41. Jayawardena R, Ranasinghe P, Byrne NM, Soares MJ, Katulanda P, Hills AP (2012) Prevalence and trends of the diabetes epidemic in South Asia: a systematic review and meta-analysis. BMC Public Health 12:380

42. Saeedi P, Salpea P, Karuranga S, Petersohn I, Malanda B, Gregg EW, Unwin N, Wild SH, Williams R (2020) Mortality attributable to diabetes in 20-79 years old adults, 2019 estimates: Results from the International Diabetes Federation Diabetes Atlas, 9(th) edition. Diabetes Res Clin Pract 162:108086

43. Mills KT, Stefanescu A, He J (2020) The global epidemiology of hypertension. Nat Rev Nephrol 16:223–237

44. Gobezie MY, Hassen M, Tesfaye NA, Solomon T, Demessie MB, Fentie Wendie T, Tadesse G, Kassa TD, Berhe FT (2024) Prevalence of uncontrolled hypertension and contributing factors in Ethiopia: a systematic review and meta-analysis. Front Cardiovasc Med 11:1335823

45. Riaz M, Shah G, Asif M, Shah A, Adhikari K, Abu-Shaheen A (2021) Factors associated with hypertension in Pakistan: A systematic review and meta-analysis. PLoS One 16:e0246085

46. Zeru MA, Tesfa E, Mitiku AA, Seyoum A, Bokoro TA (2021) Prevalence and risk factors of type-2 diabetes mellitus in Ethiopia: systematic review and meta-analysis. Sci Rep 11:21733

47. Phan DH, Vu TT, Doan VT, Le TQ, Nguyen TD, Van Hoang M (2022) Assessment of the risk factors associated with type 2 diabetes and prediabetes mellitus: A national survey in Vietnam. Medicine (Baltimore) 101:e31149

48. Kearney PM, Whelton M, Reynolds K, Muntner P, Whelton PK, He J (2005) Global burden of hypertension: analysis of worldwide data. Lancet 365:217–223

49. Defianna SR, Santosa A, Probandari A, Dewi FST (2021) Gender differences in prevalence and risk factors for hypertension among adult populations: a cross-sectional study in Indonesia. Int J Environ Res Public Health 18:6259

50. Cherfan M, Vallée A, Kab S, Salameh P, Goldberg M, Zins M, Blacher J (2020) Unhealthy behaviors and risk of uncontrolled hypertension among treated individuals-The CONSTANCES population-based study. Sci Rep 10:1925

51. Li K, Ma X, Yuan L, Ma J (2022) Age differences in the association between marital status and hypertension: a population-based study. J Hum Hypertens 36:670–680

52. Ford KJ, Robitaille A (2023) How sweet is your love? Disentangling the role of marital status and quality on average glycemic levels among adults 50 years and older in the English Longitudinal Study of Ageing. BMJ Open Diabetes Res Care 11:e003080

53. Shihab HM, Meoni LA, Chu AY, Wang N-Y, Ford DE, Liang K-Y, Gallo JJ, Klag MJ (2012) Body mass index and risk of incident hypertension over the life course: the Johns Hopkins Precursors Study. Circulation 126:2983–2989

54. Landi F, Calvani R, Picca A, Tosato M, Martone AM, Ortolani E, Sisto A, D’Angelo E, Serafini E, Desideri G (2018) Body mass index is strongly associated with hypertension: Results from the longevity check-up 7+ study. Nutrients 10:1976

55. Tang N, Ma J, Tao R, Chen Z, Yang Y, He Q, Lv Y, Lan Z, Zhou J (2022) The effects of the interaction between BMI and dyslipidemia on hypertension in adults. Sci Rep 12:927

56. Caillon A, Paradis P, Schiffrin EL (2019) Role of immune cells in hypertension. Br J Pharmacol 176:1818–1828

57. Stelmach-Mardas M, Walkowiak J (2016) Dietary Interventions and Changes in Cardio-Metabolic Parameters in Metabolically Healthy Obese Subjects: A Systematic Review with Meta-Analysis. Nutrients. 10.3390/nu8080455

58. Li Y, Jiang Y, Lin J, Wang D, Wang C, Wang F (2022) Prevalence and associated factors of diabetes mellitus among individuals aged 18 years and above in Xiaoshan District, China, 2018: a community-based cross-sectional study. BMJ Open 12:e049754

59. Saijo Y, Okada H, Hamaguchi M, Habu M, Kurogi K, Murata H, Ito M, Fukui M (2022) The risk factors for development of type 2 diabetes: Panasonic cohort study 4. Int J Environ Res Public Health 19:571

60. Campbell PJ, Carlson MG (1993) Impact of obesity on insulin action in NIDDM. Diabetes 42:405–410

61. Deng Y, Scherer PE (2010) Adipokines as novel biomarkers and regulators of the metabolic syndrome. Ann N Y Acad Sci 1212:E1–E19

62. Lorenzo M, Fernández-Veledo S, Vila-Bedmar R, Garcia-Guerra L, De Alvaro C, Nieto-Vazquez I (2008) Insulin resistance induced by tumor necrosis factor-alpha in myocytes and brown adipocytes. J Anim Sci 86:E94–104

63. Kim M-J, Lim N-K, Choi S-J, Park H-Y (2015) Hypertension is an independent risk factor for type 2 diabetes: the Korean genome and epidemiology study. Hypertens Res 38:783–789

64. Śliwińska-Mossoń M, Milnerowicz H (2017) The impact of smoking on the development of diabetes and its complications. Diabetes Vasc Dis Res 14:265–276

65. Filippini T, Malavolti M, Whelton PK, Vinceti M (2022) Sodium intake and risk of hypertension: A systematic review and dose–response meta-analysis of observational cohort studies. Curr Hypertens Rep 24:133–144

66. Grillo A, Salvi L, Coruzzi P, Salvi P, Parati G (2019) Sodium intake and hypertension. Nutrients 11:1970

67. Rust P, Ekmekcioglu C (2017) Impact of salt intake on the pathogenesis and treatment of hypertension. Hypertens from basic Res to Clin Pract 61–84

68. Kulkarni S, O’Farrell I, Erasi M, Kochar MS (1998) Stress and hypertension. WMJ 97:34–38

69. Bi Y, Jiang Y, He J, et al (2015) Status of cardiovascular health in Chinese adults. J Am Coll Cardiol 65:1013–1025

70. Yang W, Lu J, Weng J, et al (2010) Prevalence of diabetes among men and women in China. N Engl J Med 362:1090–1101

71. Vellakkal S, Subramanian S V, Millett C, Basu S, Stuckler D, Ebrahim S (2013) Socioeconomic inequalities in non-communicable diseases prevalence in India: disparities between self-reported diagnoses and standardized measures. PLoS One 8:e68219

72. Safarpour AR, Fattahi MR, Niknam R, et al (2022) Alarm of non-communicable disease in Iran: Kavar cohort profile, baseline and 18-month follow up results from a prospective population-based study in urban area. PLoS One 17:e0260227

73. Simmons SS, Hagan JEJ, Schack T (2021) The Influence of Anthropometric Indices and Intermediary Determinants of Hypertension in Bangladesh. Int J Environ Res Public Health. 10.3390/ijerph18115646

74. Kiani FZ, Ahmadi A, Babadi AS, Rouhi H (2021) Profile and preliminary results of Iranian sub cohort chronic obstructive pulmonary disease (COPD) in Shahrekord PERSIAN cohort in southwest Iran. BMC Pulm Med 21:105

75. Khoo YY, Farid NDN, Choo WY, Omar A (2022) Prevalence, awareness, treatment and control of young-onset hypertension in Malaysia, 2006–2015. J Hum Hypertens 36:106–116

76. Okello S, Muhihi A, Mohamed SF, Ameh S, Ochimana C, Oluwasanu AO, Bolarinwa OA, Sewankambo N, Danaei G (2020) Hypertension prevalence, awareness, treatment, and control and predicted 10-year CVD risk: a cross-sectional study of seven communities in East and West Africa (SevenCEWA). BMC Public Health 20:1706

77. Alsaud W, Tabbaa MJ, Kasabri VN, Suyagh MF, Abu Alsamen MA, Haddad HM, ALshweki AO (2020) Prevalence of Cardiovascular Diseases Risk Factors among Jordanians. J Saudi Hear Assoc 32:324–333

78. Odili AN, Chori BS, Danladi B, et al (2020) Prevalence, Awareness, Treatment and Control of Hypertension in Nigeria: Data from a Nationwide Survey 2017. Glob Heart 15:47

79. Mirzaei M, Mirzaei M, Sarsangi AR, Bagheri N (2020) Prevalence of modifiable cardiovascular risk factors in Yazd inner-city municipalities. BMC Public Health 20:134

80. Latt T-S, Zaw K-K, Ko K, Hlaing M-M, Ohnmar M, Oo E-S, Thein K-M-M, Yuasa M (2019) Measurement of diabetes, prediabetes and their associated risk factors in Myanmar 2014. Diabetes Metab Syndr Obes 12:291–298

81. Peltzer K, Pengpid S (2018) The Prevalence and Social Determinants of Hypertension among Adults in Indonesia: A Cross-Sectional Population-Based National Survey. Int J Hypertens 2018:5610725

82. Seclen SN, Rosas ME, Arias AJ, Huayta E, Medina CA (2015) Prevalence of diabetes and impaired fasting glucose in Peru: report from PERUDIAB, a national urban population-based longitudinal study. BMJ Open Diabetes Res & Care 3:e000110

83. Shah A, Afzal M (2013) Prevalence of diabetes and hypertension and association with various risk factors among different Muslim populations of Manipur, India. J Diabetes Metab Disord 12:52

84. Daniel OJ, Adejumo OA, Adejumo EN, Owolabi RS, Braimoh RW (2013) Prevalence of hypertension among urban slum dwellers in Lagos, Nigeria. J Urban Health 90:1016–1025

85. Muyer MT, Muls E, Mapatano MA, et al (2012) Diabetes and intermediate hyperglycaemia in Kisantu, DR Congo: a cross-sectional prevalence study. BMJ Open 2:e001911

86. Hendriks ME, Wit FWNM, Roos MTL, et al (2012) Hypertension in sub-Saharan Africa: cross-sectional surveys in four rural and urban communities. PLoS One 7:e32638

87. Anjana RM, Deepa M, Pradeepa R (2023) The ICMR-INDIAB Study: Results from the National Study on Diabetes in India. J Indian Inst Sci 103:21–32

